# Epigenome-Wide Association Study in Asian Cohort Identifies Novel DNA Methylation Markers for Carotid Intima-Media Thickness

**DOI:** 10.1101/2025.11.21.25340774

**Authors:** Konstanze Tan, Sarah E Harris, Jane Maddock, Darwin Tay, Pritesh R Jain, The HELIOS Study Team, Shi Qi Mok, Maxime Herbrard, Ulf Schminke, Can Can Xue, Liuh Ling Goh, Theresia Mina, Alexander Teumer, Weng Khong Lim, Khai Pang Leong, Khung Keong Yeo, Ching-Yu Cheng, Xueling Sim, Lee Eng Sing, Joanna M Wardlaw, Henry Völzke, Andrew Wong, Simon R Cox, Rinkoo Dalan, Abbas Dehghan, Marie Loh

## Abstract

2.

**Background:** Carotid-intima media-thickness predicts cardiovascular events and informs mechanistic research on cardiovascular diseases (CVD). However, CVD research remains Eurocentric despite etiological differences across ancestries. Incorporating Asian populations— who face substantial CVD burden with distinct etiological landscape— can enhance our understanding of cIMT biology and subclinical processes linked to CVD. This study aimed to elucidate methylation-based mechanisms of cIMT through DNA methylation profiling integrated with multi-omics data and clinically informative cIMT thresholds, leveraging an Asian cohort to enhance discovery.

**Methods:** We conducted an epigenome-wide association study (EWAS) of cIMT using peripheral blood DNA methylation at ∼850,000 CpG sites in the Asian Health for Life in Singapore (HELIOS) cohort (n=1,357), followed by targeted trans-ancestry meta-analysis with European cohorts (overall n=2,765). Causal inference analyses (summary data-based Mendelian Randomisation [SMR] and colocalisation) evaluated methylation-mediated effects on cIMT, CVD and proximal gene expression. We derived a methylation risk score (MRS) and tested its association with cIMT thresholds indicative of elevated cardiovascular risk (≥75^th^ percentile for age, sex and ethnicity).

**Results:** Three novel CpG-cIMT associations were identified (P<9.35E-07). Causal analyses supported cg08227773 methylation-mediated effects on both coronary artery disease risk (P_SMR_=2.91E-05, coloc PP.H4 =0.91) and *NBEAL2* (Neurobeachin-like 2) expression (P_SMR_=9.13E-08, coloc PP.H4=0.69), a gene implicated in immune dysregulation. MRS of cIMT aggregating the three sentinel CpGs was associated with clinically-informative cIMT elevation (Odds Ratio=2.75 for Q4 vs Q1, 95% CI: 1.47-5.13).

**Conclusions:** Through Asian-led discovery, this study identifies three novel DNA methylation markers for cIMT that are linked to cIMT elevation above clinically meaningful risk thresholds. Causal inference analyses suggest methylation-mediated CAD risk via *NBEAL2* regulation, nominating biologically relevant targets while underscoring the need for larger multi-omics resources to refine mechanisms.

## 3. INTRODUCTION

Cardiovascular disease (CVD) is the leading global cause of death and disability-adjusted life years.^1^ Preceding the overt manifestation of clinical CVD events in adulthood are lifelong atherogenic changes in youth. ^2^ Correspondingly, carotid intima media thickness (cIMT), a non-invasive ultrasound measure of atherosclerotic burden reflecting cumulative arterial remodeling, allows CVD to be studied in the subclinical phase. ^3-6^ cIMT robustly predicts incident myocardial infarction and stroke, correlates with established CVD risk factors, and also shows genetic correlation with CVD outcomes.^7-12^ However, ancestry-related differences in effect sizes of risk-factor associations and modest SNP-heritability estimates (26%-28%), point to the contribution of additional regulatory mechanisms of cIMT.^9,10^

DNA methylation— the covalent addition of a methyl group to the 5’ carbon of cytosine in CpG dinucleotides— integrates cumulative inherited and environmental exposures, representing a practical molecular tool to interrogate cIMT biology.^13^ The first published epigenome-wide association study (EWAS) of cIMT, a European multi-cohort meta-analysis (8 cohorts; n = 6,400; 450K array), identified an epigenome-wide significant signal at the canonical smoking biomarker, cg05575921 (*AHRR*) along with 33 other genomic regions.^14^ A second published EWAS of cIMT in the Dominican Republic (61 families; 769 individuals; EPIC array) reported methylation alterations at the *PM20D1* promoter, implicating mitochondrial uncoupling and reactive oxygen species generation.^15^ Crucially, cg05575921 did not replicate in the Dominican Republic study.

Motivated by the value of ancestral diversity for elucidating cIMT biology, we focus on Asian populations, whose etiologic architecture of cIMT adds complementary insight into arterial remodeling.^9,16^ We undertook a two-tier EWAS strategy: first, we meta-analysed ∼850,000 CpG across Chinese, Malays and Indian participants from the Health for Life in Singapore (HELIOS) cohort (n=1,357) for an Asian-focused screen.^17^ Signals demonstrating preliminary relevance were then advanced to trans-ancestry meta-analysis with European cohorts for validation. Robustly associated CpG sites were subsequently analysed through integrative omics approaches, then combined into a methylation risk score (MRS) for predicting clinically meaningful cIMT elevation.

## 4. METHODS

### Cohort descriptions

This section describes the Asian cohorts contributing to the cIMT EWAS and omics-based functional analyses. European cohort data were obtained from two sources: (1) pre-existing summary statistics generated from the previous published meta-analysis of cIMT (Study of Health in Pomerania [SHIP]; MRC National Survey of Health and Development [NSHD], provided by the corresponding author with permission from respective study teams, or (2) custom analysis conducted for this investigation (Lothian Birth Cohort 1936 [LBC1936]). Descriptions of the three European cohorts are provide in Supplemental Methods.

For all cohorts, participants provided written informed consent for research use of blood samples was obtained from all participants, and study protocols were approved by the relevant institutional review boards.

#### Health for Life in Singapore Study

HELIOS is a Singapore-based population cohort study designed to elucidate chronic disease determinants, enrolling 10,004 adults aged 30-84 years between 2008 and 2012, from the three major ethnic groups (Chinese, Malay and Indian). Participants comprehensive baseline assessments including clinical, lifestyle, and imaging evaluations including 3D carotid ultrasound, alongside biospecimen collection. All participants provided written informed consent for collection of questionnaire data, physical measurements, and biospecimens, plus storage of coded data/samples for future ethics-approved research. Participation was voluntary with rights to refuse any component or withdraw without penalty, and all received signed consent copies with study team and ethics office contacts. Self-reported ethnicity, used for ethnic-specific analyses in this investigation, showed high concordance with genetically determined ancestry (Chinese: 99.1%, Malay: 82.6%, Indian: 96.9%; Supplemental Methods).

HELIOS provided array-based DNA methylation data (EPIC array) from whole blood for the *de novo* cIMT EWAS, alongside whole genome sequencing (WGS) and RNA sequencing data used for eQTL and eQTM analyses, respectively (Tables S1 and S2; Supplemental Methods).

#### cIMT measurement

Bilateral cIMT measurements were obtained in the common carotid artery using the Philips iU 22 ultrasound system following established guidelines.^4,18^ Measurements were centered 1 cm proximal to the carotid bifurcation, avoiding plaque, and were taken from the lateral and posterior angles (four measurements in total for left and right cIMT). Automated measurements were obtained by using the Philips Qlab software to analyse images, with ECG gating to time the measurement at end-diastole. Manual review of automated measurements verified that the intima-media interface was correctly identified and traced in > 95% of cases. Inter-operator and inter-reader variability was consistently low (coefficient of variation <5%). cIMT_mean_ was calculated by averaging the four measurements.

#### SG10K_Health

The SG10K_Health study integrates data from five Singapore population-based adult cohorts : HELIOS, the Singapore Epidemiology of Eye Diseases (SEED) cohort, the Multi-Ethnic Cohort (MEC), the SingHealth Duke-NUS Institute of Precision Medicine (PRISM) cohort, and the TTSH Personalised Medicine Normal Controls (TTSH) cohort. ^19,20^ Four cohorts (excluding HELIOS) provided WGS and whole-blood DNA methylation profiling data (EPIC array) for *de novo* meQTL analysis to identify genetic instruments for causal inference (Table S2; Supplemental Methods). These methylation data were used exclusively for meQTL analysis and were not included in the cIMT EWAS.

### Quantification of DNA methylation

Genomic DNA was extracted from buffy coats, bisulfite-converted using the EZ DNA methylation kit (Zymo Research), and quantified using the EPIC array. Bead intensity retrieval and background correction were performed using the *minfi* R package (version 1.40.0), with detection P<0.01 was for marker-calling. Markers were excluded if non-CpG, on sex chromosomes, or if the call-rate was low (<95%). Samples were excluded based on low call-rate (<95%), mis-labelled sex, or if present in duplicate. Quantile normalisation was applied to reduce between-array technical variability. After filtering, 1,357 HELIOS samples remained for the Asian discovery EWAS (Chinese n=1,063, Malay n=150; Indian n=144). The four SG10K_Health cohorts providing methylation data for meQTL analysis underwent similar quality control procedures.

For European cohorts, whole blood-derived genomic DNA was utilised for methylation analysis using either the 450K array (LBC1936) or the EPIC array (NSHD, SHIP) ^14,21^. Quality control procedures for these cohorts were analogous to those applied in HELIOS.^14,21^

### Statistical analyses of omics data

#### Epigenome-wide association analysis

EWAS were performed separately in three HELIOS ethnic strata (Chinese, Malay and Indian; Table S1) and three European cohorts (LBC1936, SHIP, NSHD) using log-transformed cIMT_mean_ as the outcome. All EWAS regression models used broadly comparable covariates, including adjustment for age, sex, smoking status, estimated cell composition (Houseman-estimated proportions or measured counts), and technical covariates (control probe principal components or array factors) (Table _1__)._ ^22,23^

**Table 1.**
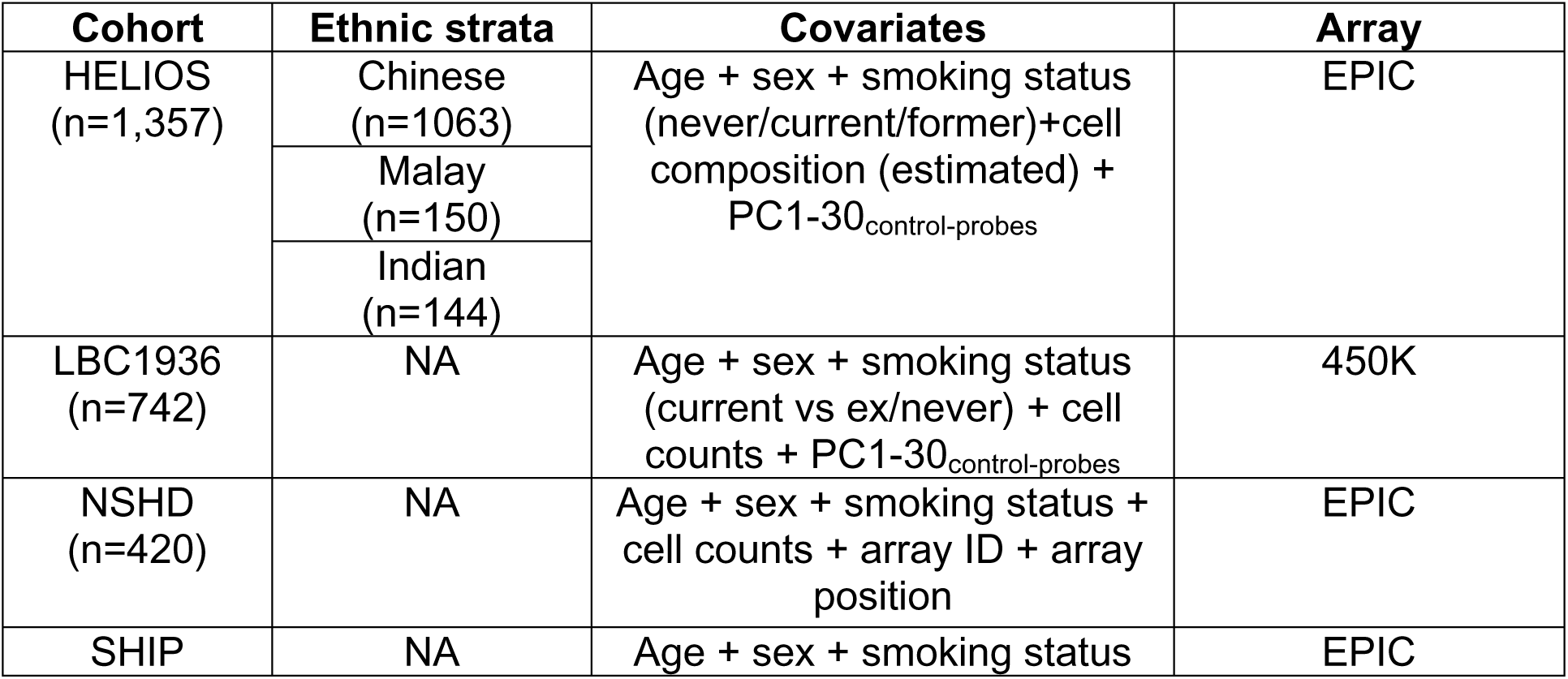
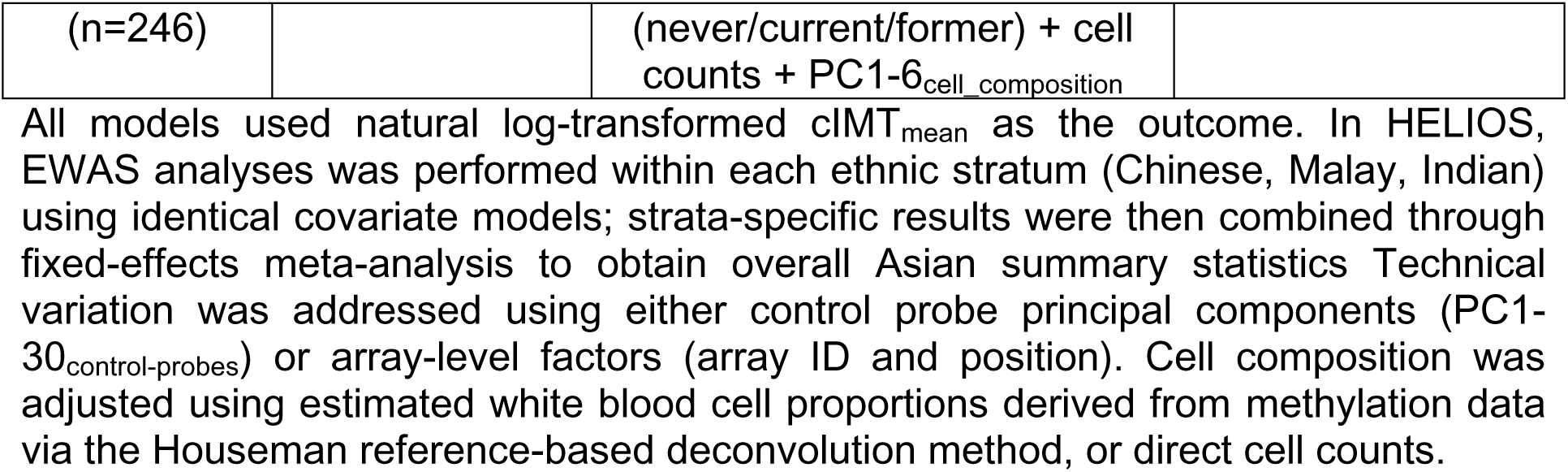
Models for epigenome-wide association study of cIMT across cohorts.

| **Cohort** | **Ethnic strata** | **Covariates** | **Array** |
| --- | --- | --- | --- |
| HELIOS<br>(n=1,357) | Chinese<br>(n=1063) | Age + sex + smoking status<br>(never/current/former)+cell<br>composition (estimated) +<br>PC1-30<sub>control-probes</sub> | EPIC |
|  | Malay<br>(n=150) |  |  |
|  | Indian<br>(n=144) |  |  |
| LBC1936<br>(n=742) | NA | Age + sex + smoking status<br>(current vs ex/never) + cell<br>counts + PC1-30<sub>control-probes</sub> | 450K |
| NSHD<br>(n=420) | NA | Age + sex + smoking status +<br>cell counts + array ID + array<br>position | EPIC |
| SHIP | NA | Age + sex + smoking status | EPIC |
| (n=246) |  | (never/current/former) + cell counts + PC1-6<sub>cell composition</sub> |  |

All models used natural log-transformed cIMT<sub>mean</sub> as the outcome. In HELIOS, EWAS analyses was performed within each ethnic stratum (Chinese, Malay, Indian) using identical covariate models; strata-specific results were then combined through fixed-effects meta-analysis to obtain overall Asian summary statistics. Technical variation was addressed using either control probe principal components (PC1-30<sub>control-probes</sub>) or array-level factors (array ID and position). Cell composition was adjusted using estimated white blood cell proportions derived from methylation data via the Houseman reference-based deconvolution method, or direct cell counts.

#### Trans-ancestry meta-analysis

A two-stage meta-analysis was conducted: Stage 1 combined three HELIOS ethnic strata for Asian-focused discovery, while Stage 2 combined Asian association statistics (CpGs with P<0.05 in Stage 1) with European results (meta-analysis of LBC1936, NSHD and SHIP) for trans-ancestry meta-analysis. Due to platform differences, LBC1936 (450K array) contributed only at overlapping 450K-EPIC sites.

All meta-analyses used the sample-size weighted Stouffer method (METAL). This approach was chosen because heterogeneity in cIMT measurement (different acquisition and summarisation protocols), as well as differences in covariate specification and sample size imbalance (Table 1) meant effect sizes may not be directly comparable, making Stouffer’s Z-based aggregation preferable to inverse variance-weighting (IVW) method for robust cross-cohort integration. ^24^

#### Expression quantitative trait methylation

Expression quantitative trait methylation (eQTM) analysis was performed in 1,201 individuals from the HELIOS cohort to identify *cis*-CpG-gene expression associations (<1Mb) (Table S2). Linear regression tested associations (MatrixEQTL R package, version 2.3), adjusting for age, sex, ethnicity, RNA Integrity number (RIN), Houseman-estimated proportions of six white blood cell subpopulations, and latent factors inferred via the Probabilistic Estimation of Expression Residuals (PEER). ^25^

#### Methylation quantitative trait loci

Methylation quantitative trait loci (meQTL) analysis was performed *de novo* in 5,273 individuals across four SG10K_Health adult cohorts to identify *cis*-SNPs associated with with sentinel CpG methylation (<1Mb), serving as instrumental variables for causal inference (Table S2). Linear regression tested associations (MatrixEQTL R package, version 2.3), adjusting for age, sex, ethnicity, Houseman-estimated proportions of six white blood cell subpopulations and methylation array control probe PCs. Cohort-specific results were combined using using fixed effects IVW meta-analysis (METAL).

#### Expression quantitative trait loci

Genome-wide expression quantitative trait loci (eQTL) analyses was performed in 1,168 individuals from the HELIOS cohort to identify *cis*-SNP-gene expression associations (<1Mb) (Table S2).^26^ Linear regression tested associations (MatrixEQTL R package, version 2.3), adjusting for age, sex, ethnicity, RIN and the top six PEER factors.

### Causal analyses

We investigated methylation-mediated effects using the Summary data-based Mendelian Randomisation (SMR) method and Bayesian genetic colocalisation (coloc R package, coloc.abf(), version 5.2.3).^27,28^ Causal inference analyses were restricted to genetic association data obtained from Asian cohorts to enhance population validity. SMR was performed using the Wald Ratio (*β̂*_*SMR*_ = *β̂*_*outcome*_ /*β̂*_*exposure*_), with *cis*-meQTLs (P<1E-05, F-statistic ≥ 10) obtained from the Asian SG10K_Health cohort as genetic instruments (Table 2). Outcome summary statistics were obtained from Asian GWAS of cIMT, CAD, ischemic stroke and myocardial infarction, and eQTL studies. Arterial eQTL data were included for exploratory analysis only.

**Table 2.** Data sources for Summary data-based Mendelian Randomisation and colocalisation analyses.

| Exposure | Exposure population (ancestry/ethnic group; N) | Outcome | Outcome data source | Outcome population (ancestry/ethnic group; N) |
| --- | --- | --- | --- | --- |
| SG10K_Health (meQTL) | Southeast Asian: Chinese, Malay, Indian, Other ethnic groups (N=5,273) | cIMT | HELIOS | Southeast Asian: Chinese, Malay, Indian ethnic groups; N=6,798 |
|  |  | Coronary artery disease | Biobank Japan<sup>29</sup> | East Asian: Japanese; N=212,453 (Cases=29,319; Controls=183,134) |
|  |  | Ischemic Stroke | Mishra et al.<sup>30</sup> | East Asian: Japanese, Korean, Chinese; N=256,274 (Cases=19,032; Controls=237,242) |
|  |  | Myocardial Infarction | Biobank Japan<sup>31</sup> | East Asian: Japanese; N=161,206 (Cases=14,992; |
|  |  |  |  | Controls=146,214) |
|  |  | Gene expression (blood) | HELIOS | Southeast Asian: Chinese, Malay Indian, Other ethnic groups; N=1,168 |
|  |  | Gene expression (aorta) | GTEx v10<sup>32</sup> | European: N=472 |
|  |  | Gene expression (coronary artery) |  | European ancestry: N=268 |
|  |  | Gene expression (tibial artery) |  | European ancestry: N=689 |

*cis*-meQTL (<1Mb) from the multi-ethnic Asian SG10K\_Health cohort were used as genetic instruments. Outcome data were obtained from GWAS of cardiovascular phenotypes (cIMT in Southeast Asian populations; coronary artery disease, ischemic stroke and myocardial infarction in East Asian populations) and eQTL analyses in blood (Southeast Asian ancestry) or arterial tissue (European ancestry). Sample sizes represent total participants, with case-control numbers specified for binary outcomes. Abbreviations: cIMT, carotid intima–media thickness; eQTL, expression quantitative trait loci analysis; GTEx, Genotype-Tissue Expression; GWAS, genome-wide association study; meQTL, methylation quantitative trait loci analysis; SMR, Summary-data Mendelian randomisation.

Colocalisation analysis was performed on ±1 Mb windows centered on each sentinel CpG site. For each region, coloc.abf() computes the posterior probabilities of five mutually exclusive hypotheses (H0-H4) describing the local genetic architecture of two traits, of which H3 and H4 indicate genetic effects for both traits within the analysed region— either via a shared (H4) or distinct causal variants (H3). We used the default priors (p₁=p₂=1E-04, p₁₂=1E-05) and performed sensitivity analyses by varying the shared-causality prior (p_12_). We prioritise CpG-outcome pairs showing SMR association (P<0.05) and strong colocalisation support (PP.H4>0.5) as consistent with methylation-mediated effects via a shared causal variant. Pairs with elevated PP.H3 (>0.5) were also flagged for the possibility of shared regulation diluted by cross-ancestry LD differences or by the presence of multiple causal variants per trait in the region.

### Construction and assessment of a methylation risk score for cIMT

An MRS was constructed from sentinel CpGs for a subset of HELIOS participants analysed in the EWAS of cIMT (n=1,353; excluding 4 participants aged >80 years for whom normative ranges were not available). Participants were classified as having elevated cIMT if their cIMT_mean_ exceeded the age- sex- and ethnicity-specific 75^th^ percentile based on Asian normative ranges.^33^ The cohort was split 70/30 into training (n=948) and testing (n=405) sets stratified by ethnicity and cIMT to maintain balance.The MRS was defined as:

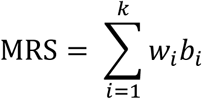

where *k* is the number of sentinel CpGs; w_i_ is the coefficient obtained from regressing sentinel CpG methylation against log-transformed cIMT_mean_ (i.e., lncIMT_mean_ ∼ CpG + Age + Sex; ethnic-stratified regression followed by sample size-weighted Stouffer meta-analysis); and b_i_ is the quantile-normalised methylation beta value. Performance in the test set (n=405) was assessed using Receiver Operating Characteristic (ROC) analysis and quartile-based logistic regression across nested models:

- Model 1 (M1): cIMT_elevated_ ∼ MRS + Age + Sex + Ethnicity
- Model 2 (M2): cIMT_elevated_ ∼ Age + Sex + Ethnicity
- Model 3 (M3) M2 + smoking

Area Under Curve (AUC) differences were tested using DeLong’s method. For quartile analysis, potential effect modification by ethnicity was evaluated by adding an MRSxEthnicity interaction term and comparison with the main-effects model via a likelihood ratio test. Robustness was assessed by re-deriving the MRS using weights from comprehensively-adjusted models (i.e., lncIMT_mean_ ∼ CpG + Age + sex + smoking status (never/current/former)+cell composition (estimated) + PC1-30_control-probes_), matching the EWAS specification.

### Gene set and transcription factor binding site enrichment analysis

Genes mapped to cIMT-linked CpGs were tested for overrepresentation in Gene Ontology (GO) biological processes and KEGG/REACTOME pathways, restricted to CpG-gene pairs with *cis*-eQTM support (P<0.05). In parallel, cIMT-associated CpGs were assessed for enrichment across 1,210 transcription factor binding sites (TFBS). Gene annotations were obtained from org.Hs.eg.db (v3.18.0); gene sets from msigdbr (v7.5.1); and TFBS data from ReMap 2022 ^34,35^.

To address sampling bias on the EPIC array, which preferentially measures CpGs near well-annotated genes and regulatory regions with distinct methylation characteristics from CpGs in other genomic regions, a permutation-based background matching strategy was used.^36,37^ An adaptive sliding-window approach was applied to select background CpGs were located >5-kb distance away from each cIMT-associated CpG, with gradually expanding matching thresholds (from ±0.025 to ±0.25 for mean methylation *β*-value; and ±0.0025 to ±0.025 for standard deviation), until 200 matches were found for each CpG. Significant enrichment was determined at P<0.005 (i.e., none of the 200 backgrounds sets equaling or exceeding test set overlap).

## 5. RESULTS

### Study design

Figure 1 outlines the analytical workflow. Briefly, Asian-focused screening followed by trans-ancestry meta-analysis with European cohorts was conducted to identify cIMT-associated CpG sites. A tiered inferential strategy was adopted: a suggestive threshold (P<1E-03) was applied to identify an inclusive set of CpGs for functional enrichment analyses, while a subset Bonferroni-corrected threshold (P<9.35E-07) was applied to designate independent, robust associations (‘Sentinel CpGs’) for targeted multi-omics and causal inference analyses, including SMR and colocalisation. We triangulated evidence from these analyses to prioritise sentinel CpGs with putative methylation-mediated involvement in cIMT, CVD and proximal gene expression (<1Mb between CpG and gene). Finally, robustly-associated CpG sites were aggregated into an MRS and evaluated against population-tailored thresholds informative for increased cardiovascular risk.

**Figure 1:**
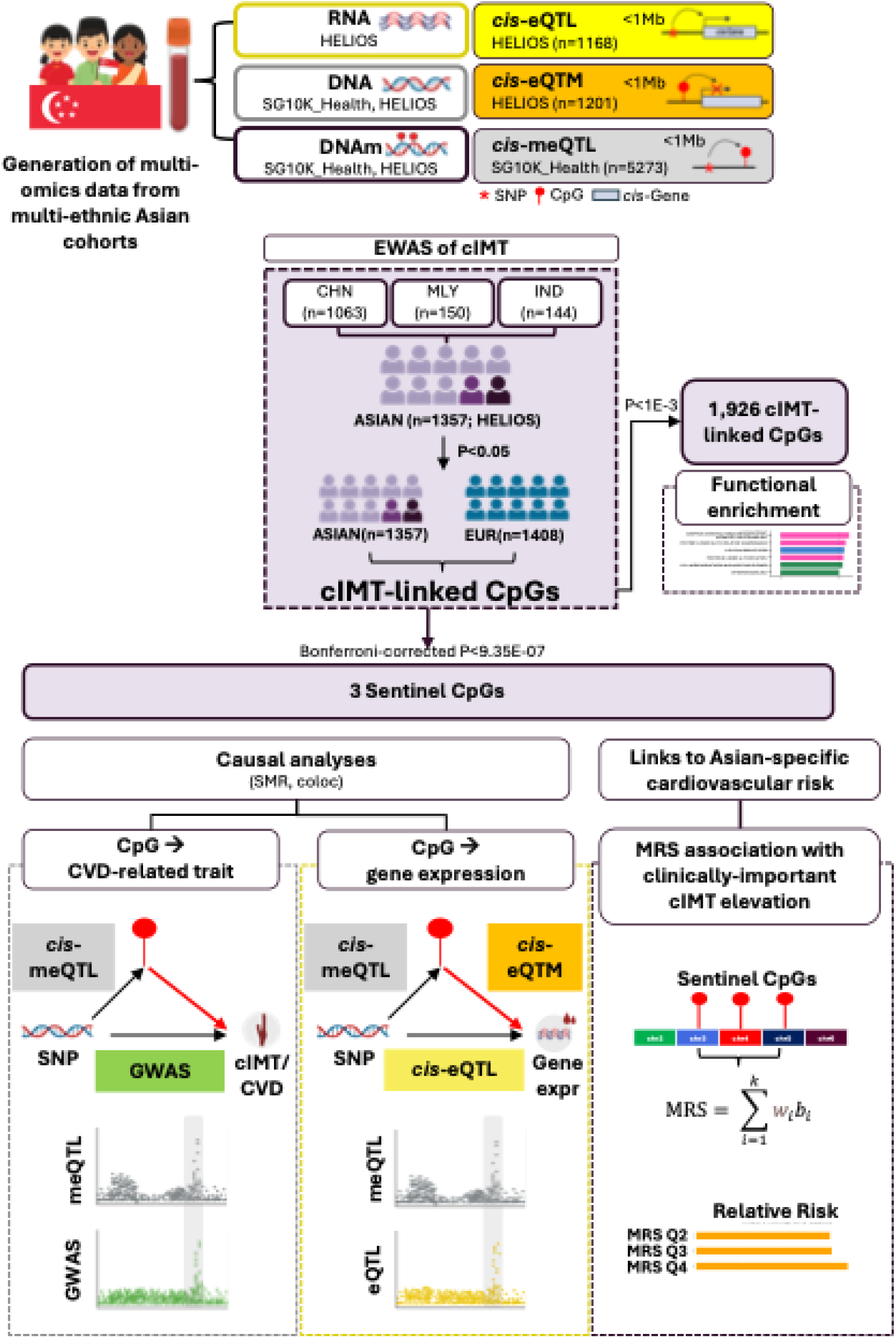
Study design. The analytical workflow integrates Asian-focused discovery with trans-ancestry validation. The Asian-focused screening stage conducted meta-analysis across ∼850,000 CpG sites across three major Asian ethnic groups—Chinese, Malay, and Indian participants—from the HELIOS cohort (n=1,357). Signals reaching P<0.05 were advanced to trans-ancestry meta-analysis with European cohorts for validation. A Bonferroni-corrected significance threshold (P<9.35E-07, 0.05/53,470 CpGs tested in trans-ancestry stage) was applied to select three robust associations (‘Sentinel CpGs’) for targeted interrogation through multi-omics integration (meQTL, eQTL, eQTM analyses) and causal inference methods to examine evidence for methylation-mediated effects on cIMT, CVD and gene expression. An MRS was constructed from the three validated methylation markers and evaluated for its association with cIMT elevation above the 75^th^ percentile based on Asian normative ranges. Abbreviations: CHN, Chinese; cIMT, carotid intima-media thickness; DNAm, DNA methylation; eQTL, expression quantitative trait loci analysis; eQTM, expression quantitative trait methylation analysis; IND, Indian; meQTL, methylation quantitative trait loci analysis; MLY, Malay; MRS, methylation risk score.

#### Epigenome-wide association study of carotid intima-media thickness

We identified CpG sites with cIMT-associated methylation changes using EWAS implemented within a two-stage meta-analytical framework. Stage 1 meta-analysed three distinct Asian ethnic strata (Chinese, Malay and Indian) from the HELIOS cohort to prioritise Asian-relevant signals, identifying 54,617 CpG sites at nominal significance (P<0.05) (Table S1, Figure S1). To reduce the multiple testing burden, the trans-ancestry meta-analysis in Stage 2 focused on CpGs which reached nominal significance in Stage 1. Methylation data were available for 53,470 of the 54,617 Stage 1-significant CpG sites (98%) across all Asian and European datasets and were included in the trans-ancestry meta-analysis (Figure 2a). Three CpGs were confirmed by the trans-ancestry meta-analysis at a subset Bonferroni-corrected significance threshold of P<9.35E-07 (0.05/53,470 tests): cg08227773 (P=3.33E-07; chr3, *CCDC12* intron), cg10556813 (P=3.37E-07; chr5, *KIF3A* intron) and cg14978069 (P=5.36E-07; chr4, *SCLT1* intron) (Figure 2a-d). Since these CpGs are located on different chromosomes, they represent independent association signals, hereafter referred to as ‘sentinel CpGs’. Sentinel CpG associations remained significant after adjusting for ethnic-specific genetic principal components (Bonferroni-corrected P<0.05/3), confirming robustness to population substructure (Table S3). Additionally, all sentinel CpG associations retained directional consistency in a subset of participants (n=837 out of 1,357) who did not have diagnosed diabetes, hypertension, hyperlipidaemia, were not on medication for these conditions, and showed no evidence of metabolic syndrome. (Table S4; Supplemental Methods).

**Figure 2:**
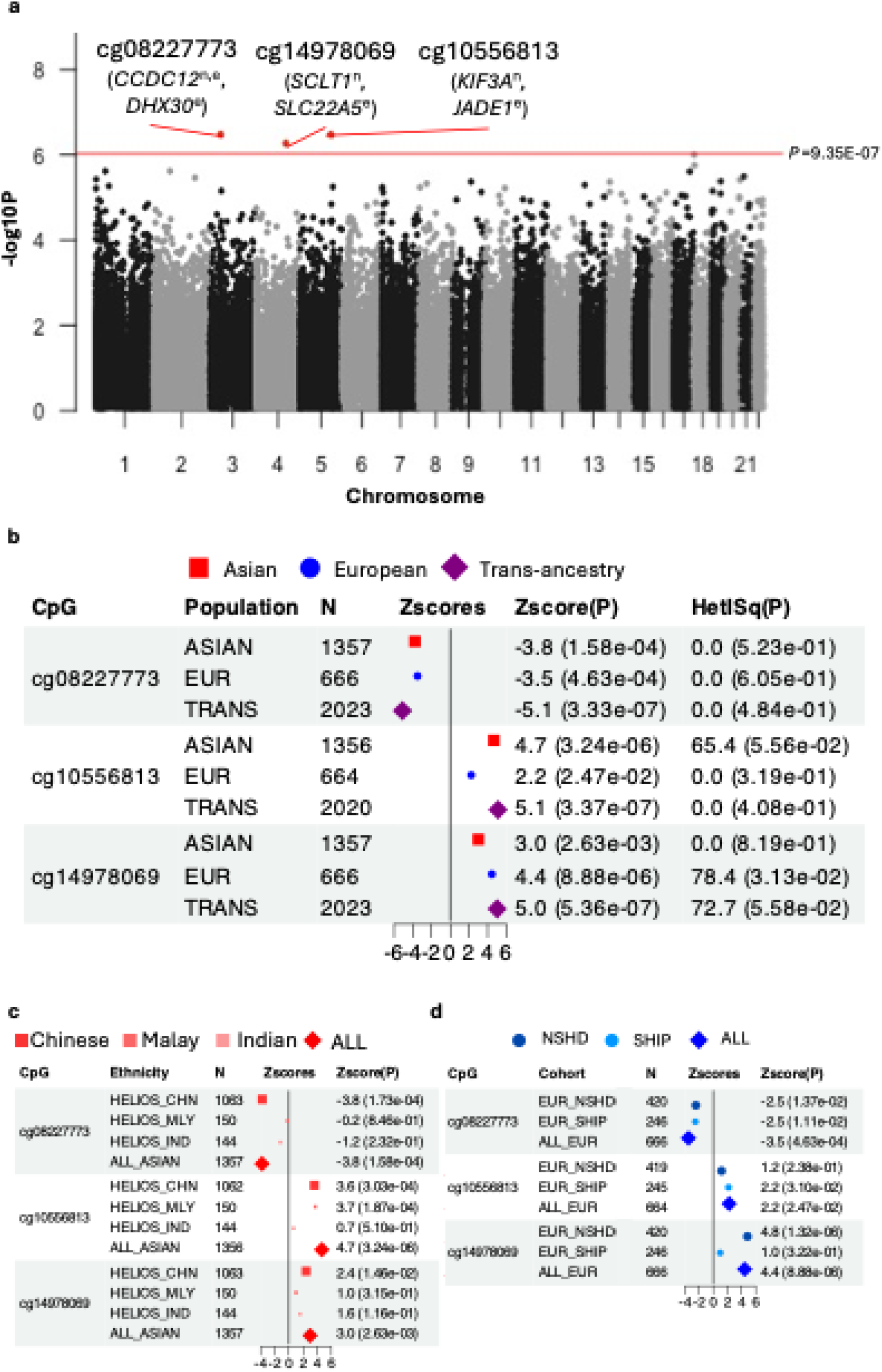
Associations between sentinel CpG sites and cIMT— (a) Manhattan plot of cIMT associations. Associations for the 53,470 CpGs included in the targeted trans-ancestry meta-analysis are shown. Sentinel CpGs (Bonferroni-corrected P<9.35E-07) are highlighted and annotated with proximal: genes: nearest gene (n) and top *cis*-eQTM-associated gene (e; P_eQTM_ <0.05). **(b-d) Forest plots of sentinel CpG associations in (b) trans-ancestry, (c) Asian-specific, and (d) European-specific meta-analyses, respectively.** For the European-specific meta-analysis, LBC1936 cohort (measured on 450K array) was not included since all sentinel CpGs were unique to the EPIC array. Data point sizes correspond to marker-specific effective sample sizes that are normalised to sum to one across all analysed groups. Test statistics from Stouffer’s sample-size weighted meta-analysis approach (Zscores, P) are indicated. Heterogeneity between meta-analysed groups was assessed using Cochran’s Q test and quantified by I^2^ (HetISq), with p-values indicating significance. Abbreviations: cIMT, carotid intima-media thickness; eQTM, expression quantitative trait methylation.

Sentinel CpGs were annotated by proximity (nearest gene) and lead *cis*-eQTM association (n=1,168, HELIOS, <1Mb). Nearest genes implicate roles in splicing (*CCDC12;* Coiled-Coil Domain Containing 12) and ciliary function (*KIF3A*, Kinesin Family Member 3A; *SCLT1*, Sodium Channel And Clathrin Linker 1).^38-40^ Of these, only cg0822773-*CCDC12* was further supported by detectable *cis*-eQTM association at P<0.05 (beta=-0.84, P=6.60E-03), while *SCLT1* and *KIF3A* lacked detectable *cis*-eQTM association with their respective sentinel CpGs: (cg14978069-*SCLT1*: beta=-0.19, P=2.21E-01; cg10556813-*KIF3A* not assessed due to low expression in the dataset). Lead *cis-*eQTM genes for sentinel CpGs are involved in cellular protein synthesis (*DHX30*; DExH-Box Helicase 30), cell cycle regulation (*JADE1*; Jade Family PHD Finger 1) and fatty acid oxidation (*SLC22A5*; Solute Carrier Family 22 Member 5). *DHX30* (cg08227773-*DHX30*, beta_eQTM_=-0.84, P_eQTM_=2.05E-05) is an ATP-dependent RNA helicase that coordinates ribosome biogenesis, protein synthesis and mitochondrial metabolism and has been implicated in cancer cell survival and neurodegenerative disease. ^41,42^ *JADE1* (Jade Family PHD Finger 1; cg14978069-*JADE1*, beta_eQTM_=-0.26, P_eQTM_=4.82E-02) encodes a chromatin remodeler that regulates the cell cycle, with dysregulation linked to tumorigenesis and impaired cellular proliferation or differentiation.^43,44^ *SLC22A5* (Solute Carrier Family 22 Member 5; cg10556813-*SLC22A5*, beta_eQTM_ =-0.25, P_eQTM_=6.09E-03) encodes a carnitine transporter; its expression is cytokine-sensitive and induced in monocyte differentiation, supporting metabolic control of immune function via carnitine dependent fatty acid oxidation.^45,46^

Querying the EWAS Catalog for prior CVD/cardiometabolic associations (P<1E-04) identified links between cg08227773 and cg14978069 and incident chronic obstructive pulmonary disease, plus cg14978069 with the inflammatory marker high-sensitivity C-reactive protein; complementary GWAS Catalog lookups of proximal genes (nearest gene and lead *cis*-eQTM) revealed that several of these loci harbor variants previously associated (P<1E-05) with multiple cardiometabolic traits, inflammatory markers, and atopic conditions (asthma, eczema), collectively implicating these sentinel CpGs and their genes in inflammation, which is central to arterial remodeling (Tables S5–S6).

#### Enhanced discovery through inclusion of Asian cohorts

The inclusion of an Asian cohort expanded the pool of candidate signals in the current EWAS. Across 53,470 CpGs analysed in the trans-ancestry meta-analysis, Asian-European concordance of test statistics was weak (Spearman ρ=0.11, p<2.2E-16), and 48,675/53,470 CpGs (91.0%) showed detectable association (P<0.05) exclusively in the Asian-focused screen, without corresponding evidence in the European-specific meta-analysis (P>0.05). This pattern is consistent with discovery gains attributable to the addition of an Asian cohort alongside a need for rigorous multiplicity control to focus on the most stable signals.

To assess the generalisability of the most robust previously reported cIMT association— cg05575921 (chr3, *AHRR* intron), which reached epigenome-wide significance in a European-only meta-analysis (discovery n=6,400; β=−0.026; P=3.5E-08)— we conducted a trans-ancestry meta-analysis at this locus. Although the association replicated in trans-ancestry meta-analysis (Z_trans_ =-3.5, P=4.07E-04) (Figure 3a), it was detected only in Europeans (Z=-3.7, P=1.80E-04) (Figures 3a,b), but attenuated in Asians (Z=-1.2, P=2.18E-01) (Figures 3a,c), possibly reflecting differences in exposure distributions.

**Figure 3:**
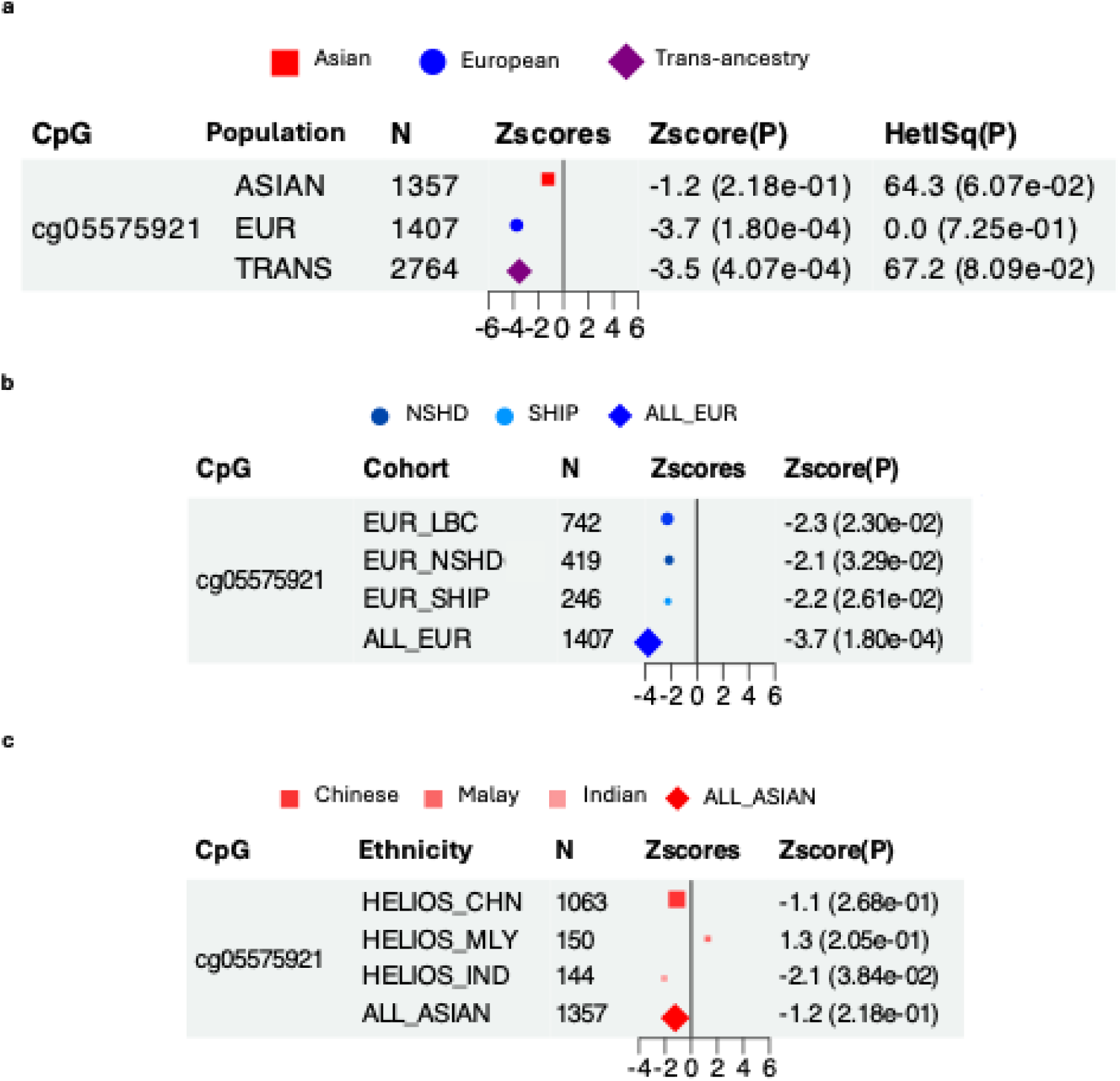
Association between cg05575921 and cIMT. The cg05575921-cIMT association was previously reported at epigenome-wide significance in a meta-analysis of European cohorts (n=6,400, Portilla-Fernández et al., 2021). **(a-c) Forest plots of the cg05575921-cIMT association** in the **(a)** trans-ancestry meta-analysis, **(b)** Asian-specific analysis and **(c)** European-specific analysis from the current investigation. The association reached P<0.05 only in the European-specific analysis, but not in the Asian-specific analysis. Data point sizes correspond to normalised effective sample size. Abbreviations: cIMT, carotid intima-media thickness

#### Gene set and transcription factor binding site overrepresentation analysis

To identify biological pathways potentially affected by cIMT-associated methylation changes, we performed gene set overrepresentation analysis on an inclusive set of 1,926 CpGs (P<1E-03). Of the 1,926 CpGs, detectable *cis*-eQTM relationships (P<0.05) were identified between 1,299 CpGs and 3,339 unique genes (<1Mb). For enrichment testing, these 3,339 genes served as the test set, compared against a background derived from 200 permuted sets of EPIC array CpGs matched to the 1,926 CpGs for methylation levels and variability. Together, the blood-enriched terms suggest a systematic arterial remodeling program encompassing re-activation of developmental circuits, immune cell supply, as well as metabolic and cytoskeletal rewiring which are known to modulate leukocyte adhesion, endothelial barrier function and mechanosensing. (Figure 4)

**Figure 4:**
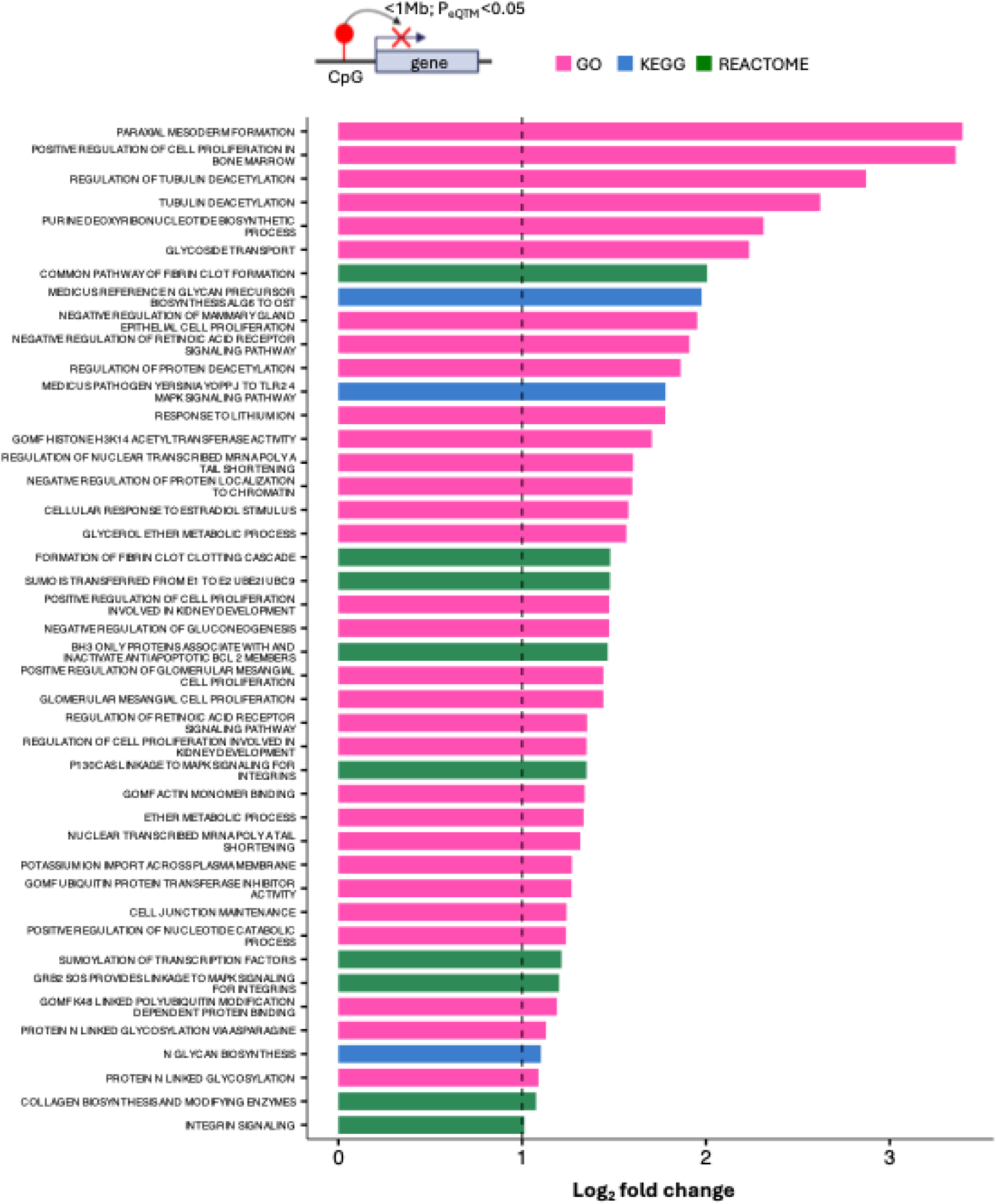
Overrepresented gene sets among genes mapped to cIMT-linked CpGs. Genes mapped to cIMT-linked CpGs were analysed for pathway enrichment against a background of genes mapped to 200 permutations of EPIC array CpGs of matched methylation level and variability to the cIMT-linked CpGs. CpG-gene mapping was done via *cis*-eQTM analysis (P_eQTM_<0.05). Enriched gene sets (P<0.005, log_2_ fold change >1) are displayed.

To further explore potential regulatory mechanisms, we tested the 1,926 cIMT-linked CpGs for enrichment in TFBS compared to matched background EPIC array CpGs. cIMT-linked CpGs were enriched for binding sites of 66 transcription factors (P <0.005), including *MEF2C* (top hit; fold-change =2.04 relative to background mean; P <0.005) and others with established roles in vascular remodeling (Table S7). Endothelial *MEF2C* deficiency disrupts atheroprotective mechanisms, increases inflammatory activation and leads to rapid vascular dysfunction *in vivo*, likely through NF-κB signaling. ^47,48^ However, Mef2c regulates macrophage polarization independently of NF-κB signaling in mice, indicating coordinate, context-dependent contributions of *MEF2C* to arterial remodeling across different cell types. ^49^

#### Genetically-anchored causal inference analyses to explore sentinel CpG-mediated pathways

Beyond observational associations, genetically-anchored causal inference analyses were applied to evaluate sentinel CpG-mediated pathways to cIMT and atherosclerotic CVD (CAD, ischemic stroke and myocardial infarction); and to putative effector genes (Table 2). Causal inference analyses were restricted to genetic association inputs from Asian cohorts to enhance population validity.

Of all CpG-trait pairs evaluated, evidence compatible with methylation-mediated involvement was observed only for cg08227773-CAD(P_SMR_ = 2.91E-05), with colocalisation indicating a 91% posterior probability that altered cg08227773 and CAD risk are driven by a shared genetic variant (coloc PP.H4=0.91) (Figure 5; Table S8). We tested how sensitive the colocalisation inference was to the assumed chance that the same variant affects both traits (p_12_). Across a wide range of plausible values (2.51E-06 to 3.16E-05, including the default 1E-05), the H4 call remained robust (coloc PP.H4>0.5 and H4/H3 ≥3), indicating stable shared-signal inference. To minimise LD mismatches from combining a multi-ethnic meQTL dataset with an East Asian-only GWAS, we repeated colocalisation analysis using *cis*-meQTL effects from the Chinese-only subset of the SG10K_Health meQTL cohort (n=2,585). Across 3,107 variants common to both datasets, the posterior for a shared causal variant remainted high (PP.H4=0.89), suggesting the colocalisation is unlikely reflect artefacts from ancestry-related LD differences.

**Figure 5:**
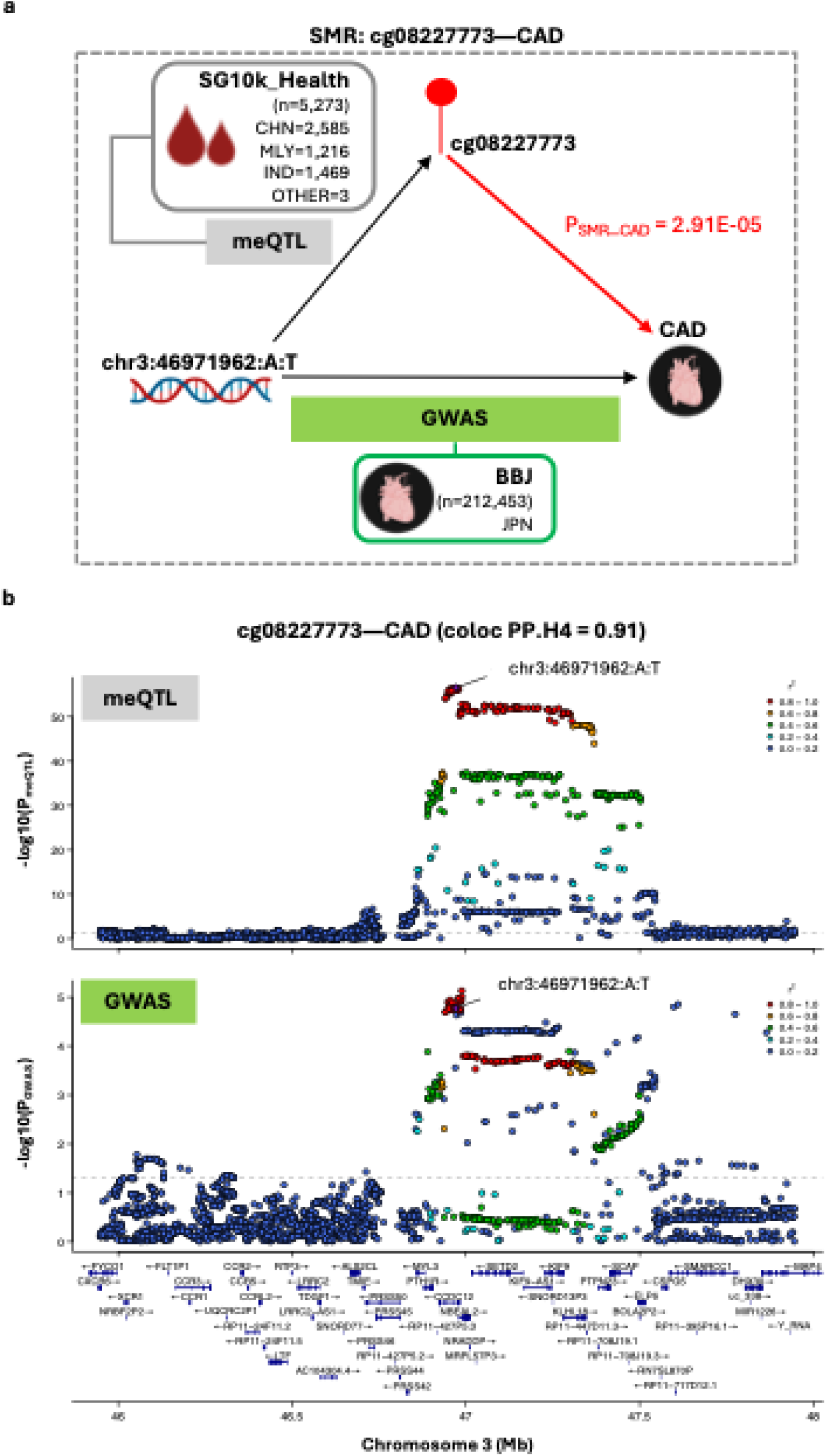
Evidence for cg08227773 methylation-mediated effects on CAD risk. **(a) Schematic overview of the SMR experiment.** *cis*-meQTL data (exposure) were derived from Asian cohorts in SG10K_Health (n=5,273). CAD GWAS summary statistics (outcome) were obtained from Biobank Japan (n=212,453 including 29,139 cases and 183,134 controls). The instrumental SNP is chr3:46971962:A:T (P_meQTL_=2.79E-57, F-statistic=306.25). **(b) Regional association plots displaying genetic associations for cg08227773 methylation and CAD.** Plots display cg08227773 meQTL (top) and CAD GWAS (bottom) signals within a ±1Mb window centered on cg08227773. The ±1Mb window size reflects the *cis*-region used for meQTL mapping in the SG10K_Health cohort. Colocalisation analysis supported shared causal variant (PP.H4=0.91). SNPs are colored according to their linkage disequilibrium (r^2^) with the instrumental SNP, calculated using the full SG10K_Health cohort with available genetic data (N=9,766). Gene tracks show locations of protein-coding genes based on Ensembl Release 86 (hg38). Abbreviations: BBJ, Biobank Japan; CAD, coronary artery disease; CHN, Chinese; IND, Indian; JPN, Japanese; MLY, Malay; meQTL, methylation quantitative trait loci analysis

Causal analyses with proximal gene expression as outcome were performed between two sentinel CpGs and 26 unique proximal genes (±1Mb) expressed in our whole blood RNAseq dataset (HELIOS, n=1,168) with suitable genetic instruments identified. Of these, evidence compatible with methylation-mediated involvement was observed only for cg08227773-*NBEAL2* (P_SMR_ = 9.13E-08), with colocalisation indicating a 69% posterior probability that altered cg08227773 methylation and *NBEAL2* expression are driven by a shared genetic variant (Figure 6; Table S9). Sensitivity analyses across plausible sharing priors showed that the H4 call remained robust over a wide range of values (1.58E-05 to 3.16E-05, including the default 1E-05), indicating a stable shared-signal inference. SMR analyses using arterial eQTL data suggest that the blood-supported cg08227773-*NBEAL2* relationship may generalise to arterial contexts (aorta P_SMR_=1.75E-04; coronary artery P_SMR_=1.36E-04;, and tibial artery P_SMR_= 2.29E-12; each instrumented by the same *cis*-variant at chr3:46971962:A:T), although interpretation remains preliminary given the mismatch in both ancestral and tissue contexts of the genetic association data (Asian blood-based meQTL and European arterial eQTL; Table 2) which may reduce power and introduce bias. Mechanistically, *NBEAL2* (Neurobeachin-like 2) is required for platelet α-granule biogenesis; loss-of function mutations in *NBEAL2* causes Grey Platelet Syndrome, an inherited bleeding disorder ^50-52^. *NBEAL2* deficiency is also linked to immune dysregulation, characterised by heightened expression of activation and degranulation markers on neutrophils and eosinophils, as well as elevated proportions of specific adaptive immune lymphocytes.^53^ Together, these findings support biological plausibility for a cg08227773–*NBEAL2* regulatory axis in arterial remodeling.

**Figure 6:**
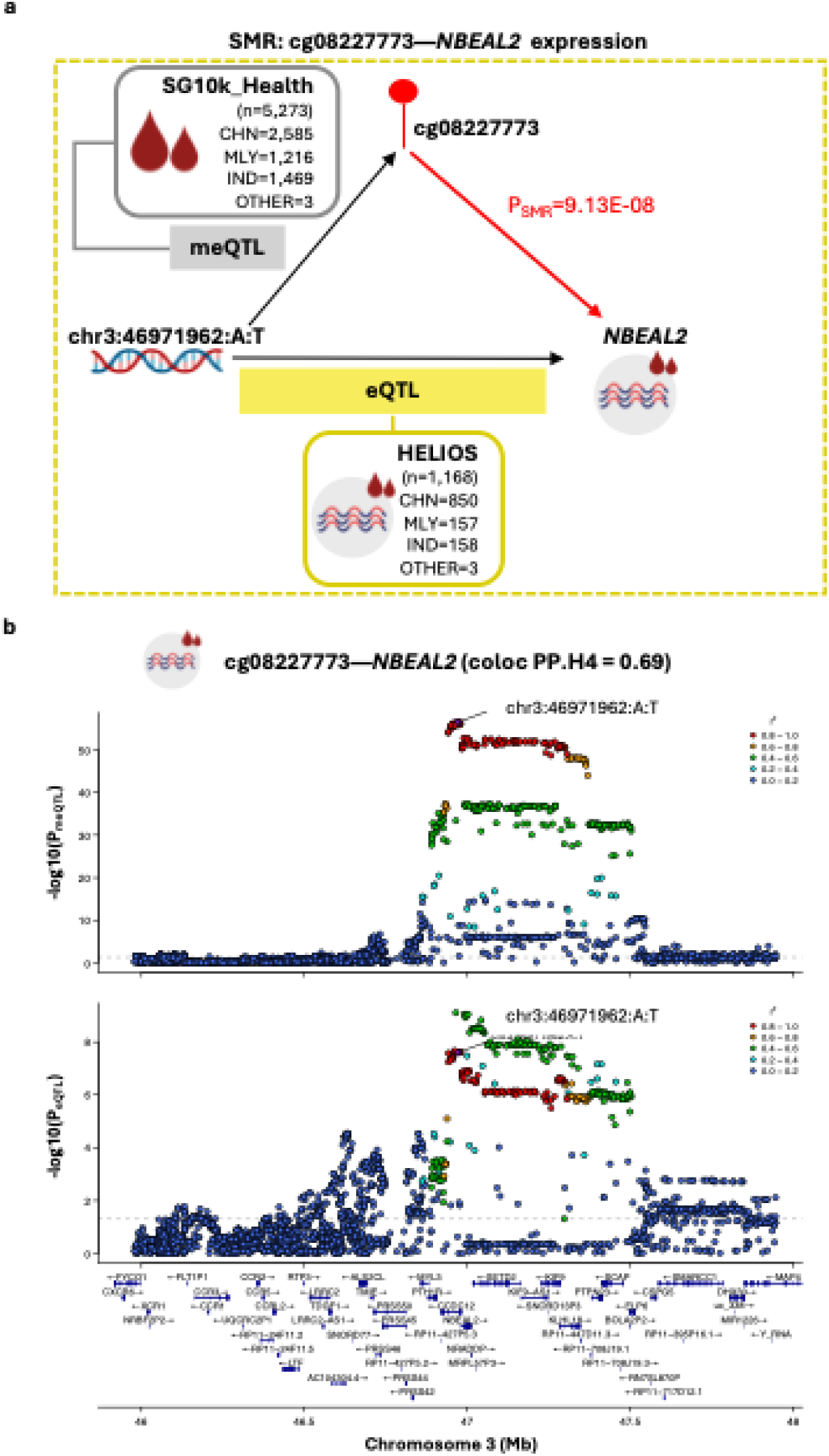
Evidence for cg08227773 methylation-mediated effects on *NBEAL2* expression. **(A) Schematic overview of the SMR experiment.** *cis*-meQTL data (exposure) were derived from Asian cohorts in SG10K_Health (n=5,273). Blood-based *cis*-eQTL data (outcome) were obtained from the Asian HELIOS cohort (n=1,168) for primary analyses. The instrumental SNP for SMR is chr3:46971962:A:T (P_meQTL_=2.79E-57, F-statistic=306.25). **(B) Regional association plots displaying genetic associations for cg08227773 methylation and *NBEAL2* expression.** Plots display cg08227773 meQTL (top) and *NBEAL2* eQTL (bottom) signals within a ±1Mb window centered on cg0822773. The ±1Mb window size reflects the *cis*-region used for meQTL mapping in the SG10K_Health cohort. Colocalisation analysis supported a shared causal variant (PP.H4=0.69). SNPs are colored according to their linkage disequilibrium (r^2^) with the instrumental SNP, calculated using the full SG10K_Health cohort with available genetic data (N=9,766). Gene tracks show locations of protein-coding genes based on Ensembl Release 86 (hg38). Abbreviations: CHN, Chinese; IND, Indian; MLY, Malay.

Apart from cg08227773-*NBEAL2,* causal inference analyses further identified 16 CpG-gene pairs for which the analysed genomic window shows effects for both methylation and expression, but driven by distinct variants (P_SMR___expr_ <0.05, coloc PP.H3 >0.5) (Table S9). While high PP.H3 formally indicates distinct variants under coloc’s single-variant model, elevated PP.H3 can also arise under truly shared-variant scenarios— particularly in multi-ethnic analyses, where cross-dataset LD differences reduce concordance, or when multiple causal variants per trait dilute an apparent single-variant colocalization signal. These 16 pairs, prioritised for future fine-mapping to resolve locus architecture, implicate genes relevant to arterial remodeling across leukocyte trafficking, inflammation and platelet function, as well as broader roles in cytoskeletal dynamics, mechanosensing and transcriptional control (Table S9).

#### Development and validation of a methylation risk score for clinically meaningful risk elevation

To assess clinical relevance of the methylation perturbations, we combined the three sentinel CpGs into a weighted MRS evaluated its relationship with clinically meaningful cIMT elevation—defined as cIMT at or above the age-, sex- and ethnicity- specific 75^th^ percentile, a threshold defined by the 2008 American Society of Echocardiography guidelines and reinforced by contemporary evidence of higher CVD event rates amongst individuals at or above this threshold ^4,54^. We used a 70:30 train-test split in the HELIOS cohort (N=1,353), stratified by ethnicity and the prevalence of at-risk cIMT, to fit the MRS in the training set (n=948) and evaluate performance in the testing set (n=405). The MRS was constructed using CpG weights estimated in models adjusted for age, sex, and ethnicity; covariates were limited to these basics to mitigate overfitting given the small sample size.

In this Asian cohort, MRS differed significantly between people with elevated cIMT and those with normal cIMT (P=1.49E-03, Figure 7a). Ethnic-specific MRS distributions did not differ detectably, indicating limited evidence for the influence of ethnic-related heterogeneity on the observed separation at the current sample sizes (Figure S2a). Across ethnic strata, the MRS distribution was shifted toward higher values among individuals with above-threshold cIMT; however the separation only reached P<0.05 in the Chinese ethnic strata (n=318), possibly reflecting limited power in the Malay (n=44) and Indian (n=43) samples (Figure S2b).

**Figure 7.**
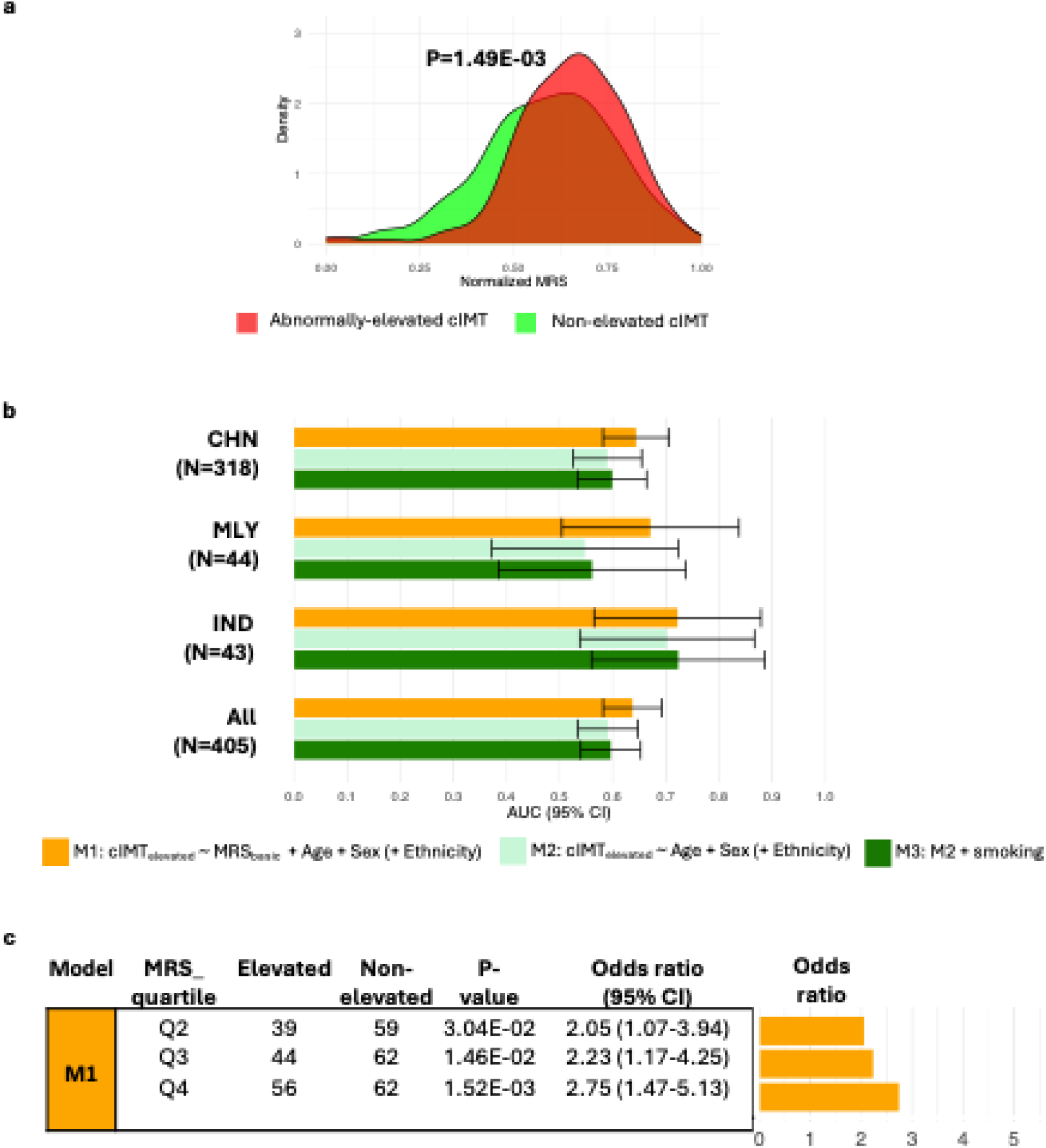
MRS of cIMT predicts clinically meaningful cIMT elevation. The MRS was built from sentinel CpGs, weighting each CpG by its Stouffer-combined z-score estimated in the training set (model: log-transformed cIMT_mean_ ∼sentinel CpG methylation + age + sex). Models were fit separately by ethnicity, and the per-ethnicity association z-scores were combined using Stouffer’s method to obtain the final CpG weight. **(a) Distribution of MRS by cIMT status.** MRS values were normalised within each group for visualisation purposes. P-values are from two-sample t-tests using the original (unnormalised) scores. **(b) Discriminatory ability of MRS.** ROC analysis compares models with and without the MRS; AUCs are plotted **(c) Stratification by MRS quartiles.** Participants were grouped by MRS quartile; relative odds of clinically concerning cIMT elevation are reported relative to Q1 (lowest MRS). Odds ratios and 95% CI were obtained from logistic regression (cIMT_elevated_ ∼ MRS quartile + Age + Sex + Ethnicity) by exponentiating the MRS quartile coefficients. Abbreviations: AUC, Area Under Curve; CHN, Chinese; IND, Indian; MLY, Malay; MRS, methylation risk score; ROC, Receiver Operating Characteristic

Receiver Operating Characteristic (ROC) analysis across three stepwise models (M1-M3) showed that the MRS provides moderate discrimination for above-threshold cIMT (Figure 7b; Table S10). There were no detectable differences in MRS discriminatory ability (AUC for M1) across ethnic strata (pairwise DeLong tests for AUC differences: all P>0.05). In the overall multi-ethnic cohort, adding the MRS to a demographics-only baseline (M2 versus M1) increased Area Under Curve (AUC) by 7.72% (AUC=0.59 to AUC=0.64; P=2.79E-02) (Table S10). These findings suggest that the MRS adds modest discriminatory signal for above-threshold cIMT.

Quartile analyses showed a graded association between MRS and above-threshold cIMT, with individuals at the highest MRS quartile (Q4) exhibiting 2.75-fold higher odds of abnormal cIMT elevation compared to the lowest quartile (95% CI: 1.47-5.13; P=1.52E-03, Figure 7c). Including an MRSxEthnicity interaction term in the logistic regression did not improve model fit (likelihood-ratio P>0.05), providing no detectable evidence that the association between MRS and elevated cIMT differs by ethnicity at current sample sizes.

To assess robustness to potential confounders, we derived an alternative score (MRS_comprehensive_) using CpG weights obtained from ethnicity-specific models adjusted for age, sex, smoking status, estimated white blood-cell composition and array control-probe PCs; the resulting weights were then meta-analysed across ethnicities, matching the EWAS specification. Compared to the basic score, MRS_comprehensive_ showed a larger between-group differences (above-threshold versus normative cIMT P=6.08E-04; Figure S3a versus Figure 7a), better discrimination (fully adjusted model M3 AUC=0.75 (95% CI:0.71-0.80), Figure S3b versus Figure 7b) and stronger risk stratification (Q4 versus Q1 OR=5.14, (95% CI: 2.37-11.13); Figure S3c versus Figure 7c). Although comprehensive covariate adjustment strengthens the effects, these patterns are already evident with the basic score, indicating that the observed associations are robust to covariate specification.

## 6. DISCUSSION

The current EWAS meta-analysis of cIMT leveraged a multi-ethnic Asian cohort and expanded EPIC array coverage to elucidate methylation-related pathways of arterial remodeling. To ensure capture of signals from Asians, previously noted to have unique cIMT etiology but underrepresented in molecular studies, Asian-focused discovery (n=1357) was undertaken within a multi-ethnic Asian population to prioritise signals before downstream trans-ancestry meta-analysis was applied to boost statistical power for identifying robust associations (n=2765). The identified cIMT-linked CpGs implicate coordinated vascular remodeling programs operating within broader developmental, immune, and cytoskeletal regulatory frameworks. Three sentinel CpGs representing the most robust associations surfaced by trans-ancestry meta-analysis (subset Bonferroni P<9.35E-07) underwent in-depth using population-matched resources, including well-powered genetic association data and calibrated cIMT thresholds to support valid inference on biological and clinical relevance. Causal inference analyses leveraging some of the largest Asian genetic association studies within their respective phenotypes provided evidence compatible with cg08227773 methylation-mediated effects on coronary artery disease (CAD) risk and the expression of the immune regulator *NBEAL2* driven by a shared causal variant. MRS aggregating the three sentinel CpGs examined against Asian-tailored cIMT thresholds demonstrated discriminatory ability, with odds of being at elevated cIMT tracking progressively with methylation burden. Together, these findings establish that Asian cohort inclusion yields insights relevant to arterial remodelling that may extend to overt clinical CVD.

Our trans-ancestry datasets enabled a closer assessment of whether the sole reported cIMT-associated biomarker, cg05575921, generalises across populations. This canonical smoking biomarker replicated in trans-ancestry analyses but not within the Asian-only subset; consistent with prior findings in Europeans, the overall signal was largely European-driven, with directionally consistent but inconclusive evidence in the Asian-specific analysis (P>0.05). Definitive inferences about population specificity cannot be drawn due to potential non-biological sources of heterogeneity, including differences in study settings, exposure distribution, cIMT measurement protocols, phenotypic derivation and covariate specification.

A key contribution of this work is its application of genetically-anchored causal inference methods to explore the mediatory role of sentinel CpGs, beyond observational associations. We leveraged recent, large-scale Asian GWAS datasets for cIMT (HELIOS cohort, n=6,798) and CVD (East Asian cohorts, n=161,206-256,274), and addressed the lack of comprehensive Asian QTL resources by conducting *cis*-meQTL analysis in 5,273 multi-ethnic Asian individuals (SG10K_Health) and *cis*-eQTL analysis and in a non-overlapping cohort of 1,168 multi-ethnic Asian individuals (HELIOS). While these analyses did not suggest sentinel CpG methylation-mediated effects on cIMT— possibly limited by the relatively modest GWAS sample size— our analyses provide evidence consistent with cg08227773-mediated effects on CAD risk and *NBEAL2* expression via a shared causal variant. Beyond established roles in platelet function and thrombosis, emerging descriptions of *NBEAL2*’s immunoregulatory roles in lymphocytes and neutrophils suggest broader influences on the inflammatory milieu with potential effects on arterial remodeling warranting further exploration.^53,56,57^ However, mechanistic refinement is required, as the single-instrument SMR analyses undertaken in this investigation preclude pleiotropy-robust approaches that require multiple LD-independent instruments, leaving causal direction suggested but not established. Furthermore, while the vascular relevance of cg08227773-*NBEAL2* is suggested by SMR analyses using arterial eQTL data, this finding should be considered hypothesis-generating given the tissue and population mismatch between the Asian blood-derived meQTL and European arterial eQTL data used in this exploratory analysis. Priorities for mechanistic refinement include: (i) repeating the causal inference analyses using expanded Asian meQTL resources with sufficient power within individual ethnic subsets to obtain robust genetic associations and to define LD-independent instruments; and (ii) experimental manipulation of cg08227773 methylation and *NBEAL2* expression in relevant vascular cell types to assess vascular relevance.

Beyond implicating biologically-relevant genes and pathways, the cIMT-linked methylation alterations were associated with Asian-tailored cIMT thresholds informative for cardiovascular risk, consistent with capture of disease-relevant biology. An MRS constructed from three sentinel CpGs differentiated risk groups and showed graded increase in odds of beyond-threshold cIMT elevation. However, the modest training sample and shared underlying population between training and test cohorts limit the current MRS as a standalone clinical tool. Priorities to improve clinical utility include expanding training datasets to derive robust weights, and evaluating generalisability in fully independent cohorts. To assess clinical value more definitively, MRS should be tested for association with incident atherosclerotic CVD events and for incremental predictive performance beyond cIMT and standard risk factors, leveraging biobank-scale resources integrating multi-omics, detailed phenotypes, and longitudinal outcomes (e.g. from electronic health records).

## CONCLUSION

This study identifies three novel blood-based DNA methylation markers for cIMT using an Asian-led, trans-ancestry discovery strategy; among these, cg08227773 showed causal-inference evidence consistent with methylation-mediated effects on CAD risk and with regulation of *NBEAL2* expression. MRS aggregating these three signals associated with cIMT elevation defined using Asian-specific thresholds. These findings nominate biologically relevant targets while highlighting the need for larger, population-matched multi-omics resources and longitudinal CVD outcomes to refine mechanisms and more rigorously evaluate clinical utility.

## Data Availability

The datasets used and/or analysed during the current study are available from the corresponding author on reasonable request.

## NON-STANDARD ABBREVIATIONS AND ACRONYMS

450K array: Illumina Infinium HumanMethylation450 BeadChip
CAD: Coronary artery disease
cIMT: Carotid intima-media thickness
CpG: Cytosine-phosphate-Guanine dinucleotide, referring to primary sites of DNA methylation in the genome where cytosine is followed by guanine
CVD: Cardiovascular disease
EPIC array: Illumina Infinium HumanMethylationEPIC BeadChip
eQTL: Expression quantitative trait locus (SNP—gene expression association)
eQTM: Expression quantitative trait methylation (association between CpG methylation and gene expression)
EWAS: Epigenome-wide association analysis
GWAS: Genome-wide association study
LD: Linkage disequilibrium
meQTL: Methylation quantitative trait locus (SNP—CpG methylation association)
MRS: Methylation risk score
PC: Principal components
PCA: Principal component analysis
SMR: Summary data-based Mendelian Randomisation
SNP: Single nucleotide polymorphism
TFBS: Transcription factor binding sites

## 7. ACKNOWLEDGMENTS

The authors express their gratitude to all study participants for their valuable contributions, and to past and present study team members for their dedicated work in data collection and management. We particularly thank the HELIOS Study Team, as HELIOS served as the core cohort for this study, providing the majority of data including genetic, methylation, and transcriptomic datasets. A complete list of HELIOS team members and affiliations is provided below:

## The HELIOS Study Team (names listed in alphabetical order)

Akash Bahai^1^, Benjamin Lam^2,3^, Dorrain Low^1^, Elio Riboli^4^, Gervais Wansaicheong^1,5^, Harinakshi Sanikini^3^, Hong Kiat Ng^1^, Jimmy Lee^1,6^, Joanne Ngeow^1,7^, Kelvin Li^8^, Kostas Tsilidis^4^, Nilanjana Sadhu^1^, Paul Elliott^4^, Sabrina Wong^1,9^, Terry Tong^1^, Tock Han Lim^8^, Tricia Chang^10^, Wai Kee Kok^10^, Xiaoyan Wang^1^, Yik Weng Yew^1,11^

1. Population and Global Health, Nanyang Technological University, Lee Kong Chian School of Medicine, Singapore, Singapore.

2. Nanyang Technological University, Lee Kong Chian School of Medicine, Singapore, Singapore.

3. Khoo Teck Puat Hospital, Singapore, Singapore.

4. Department of Epidemiology and Biostatistics, Imperial College London, School of Public Health, London, United Kingdom.

5. Diagnostic Radiology, Tan Tock Seng Hospital, Singapore, Singapore.

6. Research Division, Institute of Mental Health, Singapore, Singapore.

7. Division of Medical Oncology, National Cancer Centre, Singapore, Singapore.

8. Ophthalmology, Tan Tock Seng Hospital, Singapore, Singapore.

9. Clinical Research Unit, National Healthcare Group Polyclinics, Singapore, Singapore.

10. National Healthcare Group Polyclinics, Singapore, Singapore.

11. Research Division, National Skin Centre, Singapore, Singapore.

Certain figures in this publication were created using BioRender (https://BioRender.com/0q97c0o).

## 8. SOURCES OF FUNDING

For authors: SEH is supported by a National Institutes of Health (NIH) research grant (U01AG083829). JMW is part funded by the UK Dementia Research Institute Ltd which receives its funding from the UK Medica Research Council, Alzheimer’s Society and Alzheimer’s Research UK. SRC is supported by a Sir Henry Dale Fellowship jointly funded by the Wellcome Trust and the Royal Society (221890/Z/20/Z).

For the European cohorts: The UK Medical Research Council provides core funding for the MRC National Survey of Health and Development [MC_UU_00019/1]. SHIP is part of the Community Medicine Research Network of the University Medicine Greifswald, which is supported by the German Federal State of Mecklenburg-West Pomerania. LBC1936 is supported by the Biotechnology and Biological Sciences Research Council, and the Economic and Social Research Council [BB/W008793/1], Age UK (Disconnected Mind project), the Milton Damerel Trust, and the University of Edinburgh. Methylation typing for LBC1936 was supported by Centre for Cognitive Ageing and Cognitive Epidemiology (Pilot Fund award), Age UK, The Wellcome Trust Institutional Strategic Support Fund, The University of Edinburgh, and The University of Queensland.

For Asian cohorts: the HELIOS cohort (NTU IRB: 2016-11-030) is supported by Singapore Ministry of Health’s (MOH) National Medical Research Council (NMRC) under its OF-LCG funding scheme (MOH-000271 and MOH-001792) and NMRC through National Cohorts Office (P2022-02-03) and the National Precision Medicine (NPM) Programme. NPM Programme Phase II is supported by the National Research Foundation, Singapore (NRF) under the RIE2020 White Space (MOH-000588 and MOH-001264) and administered by the Singapore Ministry of Health through the National Medical Research Council (NMRC) Office, MOH Holdings Pte Ltd. NPM Programme Phase III is supported by the Singapore Ministry of Health through the NMRC Office, MOH Holdings Pte Ltd under the NMRC RIE2025 NPM Phase III Funding Initiative (MOH-001734). HELIOS is also supported by intramural funding from Nanyang Technological University, Lee Kong Chian School of Medicine and National Healthcare Group. Data for SG10K_Health cohorts were generated as part of the Singapore National Precision Medicine program funded by the Industry Alignment Fund (Pre-Positioning) (IAF-PP: H17/01/a0/007). Besides HELIOS, this study made use of data / samples collected in the following SG10K_Health cohorts in Singapore: (1) The Singapore Epidemiology of Eye Diseases (SEED) cohort at Singapore Eye Research Institute (SERI) (supported by NMRC/CIRG/1417/2015; NMRC/CIRG/1488/2018; NMRC/OFLCG/004/2018); (2) The Multi-Ethnic Cohort (MEC) cohort (supported by NMRC grant 0838/2004; BMRC grant 03/1/27/18/216; 05/1/21/19/425; 11/1/21/19/678, Ministry of Health, Singapore, National University of Singapore and National University Health System, Singapore); (3) The SingHealth Duke-NUS Institute of Precision Medicine (PRISM) cohort (supported by NMRC/CG/M006/2017_NHCS; NMRC/STaR/0011/2012, NMRC/STaR/ 0026/2015, Lee Foundation and Tanoto Foundation); (4) The TTSH Personalised Medicine Normal Controls (TTSH) cohort funded (supported by NMRC/CG12AUG17 and CGAug16M012).

## 9. DISCLOSURES

### ETHICS APPROVAL AND CONSENT TO PARTICIPATE

All participating cohorts received appropriate ethical approval from their respective institutional review boards or research ethics commitees. For European cohorts, details on the ethics approvals for the contributing cohorts are as follows: LBC1936 (Scotland A Research Ethics Committee: 07/MRE00/58; Waves 2-6); NSHD (Central Manchester Research Ethics Committee for genetic studies of lifelong health, disease and ageing (07/H1008/168) and the Scottish A Research Ethics Committee); and SHIP (Ethics Committee of the University of Greifswald, BB 39/08). For Asian cohorts, details on ethics approvals for the contributing cohorts are as follows: HELIOS (Nanyang Technological University IRB 2016-11-030); SEED (SingHealth CIRB 2018/2717, 2018/2921, 2012/487/A, 2015/2279, 2018/2006, 2018/2594, 2018/2570); MEC (National University of Singapore CIRB 13-512); PRISM (SingHealth CIRB 2013/605/C); and Tan Tock Seng Hospital Personalised Medicine Normal Controls (National Health Group approvals TB-2020-001 and BTC-2020-001).

### COMPETING INTERESTS

The authors declare that they have no competing interests

## 10. SUPPLEMENTAL MATERIAL

Supplemental Methods (‘supplemental_methods.docx’)

Tables S1-S10 (‘supplemental_tables.xlsx’)

Figure S1-S3 (‘supplemental_figures.docx’)

## Notes

### Competing Interest Statement

The authors have declared no competing interest.

### Clinical Trial

NA- not a clinical trial

### Author Declarations

ETHICS APPROVAL AND CONSENT TO PARTICIPATE All participating cohorts received appropriate ethical approval from their respective institutional review boards or research ethics commitees. For European cohorts, details on the ethics approvals for the contributing cohorts are as follows: LBC1936 (Scotland A Research Ethics Committee: 07/MRE00/58; Waves 2-6); NSHD (Central Manchester Research Ethics Committee for genetic studies of lifelong health, disease and ageing (07/H1008/168) and the Scottish A Research Ethics Committee); and SHIP (Ethics Committee of the University of Greifswald, BB 39/08). For Asian cohorts, details on ethics approvals for the contributing cohorts are as follows: HELIOS (Nanyang Technological University IRB 2016-11-030); SEED (SingHealth CIRB 2018/2717, 2018/2921, 2012/487/A, 2015/2279, 2018/2006, 2018/2594, 2018/2570); MEC (National University of Singapore CIRB 13-512); PRISM (SingHealth CIRB 2013/605/C); and Tan Tock Seng Hospital Personalised Medicine Normal Controls (National Health Group approvals TB-2020-001 and BTC-2020-001).

